# Refining the Allostatic Self-Efficacy Theory of Fatigue and Depression Using Causal Inference

**DOI:** 10.1101/2024.06.17.24309015

**Authors:** Alexander J. Hess, Dina von Werder, Olivia K. Harrison, Jakob Heinzle, Klaas Enno Stephan

## Abstract

Allostatic self-efficacy (ASE) represents a computational theory of fatigue and depression. In brief, it postulates that (i) fatigue is a feeling state triggered by a metacognitive diagnosis of loss of control over bodily states (persistently elevated interoceptive surprise); and that (ii) generalisation of low self-efficacy beliefs beyond bodily control induces depression. Here, we convert ASE theory into a structural causal model (SCM). This allows for identification of empirically testable hypotheses regarding casual relationships between variables of interest. We use conditional independence tests on questionnaire data from healthy volunteers (N=60) to identify contradictions to the proposed SCM. Moreover, we estimate two causal effects proposed by ASE theory using three different methods.

Our analyses suggest that, in healthy volunteers, the data are not fully compatible with the proposed SCM. We therefore refine the SCM and present an updated version for future research. Second, we confirm the predicted negative average causal effect from metacognition of allostatic control to fatigue across all three different methods of estimation. Our study represents an initial attempt to refine and formalise ASE theory using methods from causal inference. Our results confirm key predictions from the ASE theory but also suggest revisions which require empirical verification in future studies.

## INTRODUCTION

Fatigue is a prominent symptom of major clinical significance in numerous disorders across medical disciplines^7,44^. It is fundamentally disabling for patients and profoundly affects their quality of life^10^. Fatigue is a common feature across a wide range of immunological and endocrine disorders, cancer, and neuropsychiatric diseases. In particular, it constitutes one of the core diagnostic criteria of major depression in standard psychiatric classification schemes (ICD-10 and DSM-5;^2,22^).

The clinical concept of fatigue is a heterogeneous construct, and fatiguability of cognitive and motor processes needs to be distinguished from the subjective perception of fatigue^18^. This study focuses on the latter. The pathophysiological mechanisms leading to fatigue are likely diverse^18^. Previous theories have focused on a variety of neurophysiological, immunological and inflammatory processes. Unfortunately, there are no mechanistically interpretable clinical tests available for fatigue that could be used to guide individual treatment^18^.

More recently, a novel perspective on fatigue has been proposed – the allostatic self-efficacy theory (ASE;^18,27,40^). The ASE theory is based on computational concepts of brainbody interactions^27,40^ which, in turn, are conceptually related to and inspired by Bayesian theories of perception (predictive coding;^12^) and action (active inference;^13^). The ASE theory emphasises the role of two cognitive factors for fatigue: interoception and metacognition.

Interoception corresponds to the perception of bodily states and is of major importance for understanding determinants of mental health^15,20^. Many contemporary concepts of interoception are grounded in Bayesian theories of perception and conceptualise interoception as an inference process based on the brain’s generative model of sensory inputs from the body^1,15,27,28,36,37^. More specifically, interoception can be conceptualised as “inferences about bodily (physiological and biochemical) states that are coupled to regulatory processes which serve to control these states”^41^. Metacognition can be summarised as cognition about cognition^11^, comprising a variety of evaluation processes by which the brain monitors its own performance. Building on a generic mathematical model of brainbody interactions, the ASE theory describes how the brain attempts to control bodily states via monitoring interoceptive surprise (as an index of the degree of dyshomeostasis;^40^).

In brief, the ASE theory proposes that the subjective experience of fatigue arises when, in a situation of persistent dyshomeostasis (and thus enduringly elevated interoceptive surprise), the brain arrives at the metacognitive diagnosis that its control over bodily states is failing; a condition also referred to as low allostatic self-efficacy. (Put differently, fatigue is a feeling state signalling the imperative need to rest because regulatory actions fail to resolve dyshomeostasis.) Once a generalisation of low self-efficacy beliefs beyond the body has taken place, leading to a general sense of helplessness and perceived lack of control, this is postulated to trigger the onset of depression^33,40^.

At present, the ASE theory is arguably the only concept of fatigue that explains its ubiquitous occurrence across chronic disorders. It offers testable predictions based on either (i) computational quantities (prediction error or surprise) which can be estimated from behavioural and/or neurophysiological data or on (ii) self-report data about perceived control over bodily states (metacognition of allostatic control). In this study, we focus on the latter option.

Empirically, there is initial evidence that metacognition of allostatic control – as measured by a self-report questionnaire – is inversely associated with fatigue, as predicted by ASE theory^33^. However, a comprehensive investigation of the predictions made by the ASE theory is still lacking to date. Furthermore, as almost all disease concepts in psychiatry, ASE theory has been formulated verbally, but not as a precise causal model.

Here, we present an initial attempt to tackle the latter issue. To this end, we identify variables of central interest in the ASE theory, namely metacognition of allostatic control (*M*; specifically, the feeling of being in control over one’s own bodily states), fatigue (*F*), general self-efficacy (*S*), and depression (*D*). We then formalize the causal structure implied by the ASE theory in the language of causal inference, more precisely, in the form of a structural causal model (SCM;^5,24,25^). In contrast to classical probabilistic models, an SCM induces not only an observational distribution but also a set of so-called interventional distributions. In other words, an SCM predicts how a system reacts under interventions^43^. We make use of a publicly available empirical dataset to test key aspects of the structure of the proposed SCM. Moreover, we use established methods for the estimation of average causal effects focusing on central aspects of the ASE theory.

## MATERIALS AND METHODS

### Empirical Dataset

In this work, we used data from a previous study conducted at the Translational Neuromodeling Unit (TNU) Zurich, the perception of breathing in the human brain (PBIHB) study; a detailed description of the dataset can be found elsewhere^14^. It comprises behavioural, questionnaire and neuroimaging data from 60 healthy individuals. The questionnaire data used for our analysis are freely available for download from the Zenodo open data repository at https://doi.org/10.5281/zenodo.10992529. Participants completed a battery of psychological questionnaires assessing subjective affective measures, both general and breathing-specific subjective interoceptive beliefs as well as measures of general positive and negative affect, resilience, self-efficacy and fatigue. For our analysis, we focused on the following measures as representations of the central quantities of the ASE theory:

- **fatigue (***F*): Fatigue Severity Scale (FSS)
- **general self-efficacy (***S*): General Self-Efficacy Scale (GSES)
- **depression (***D*): Centre for Epidemiologic Studies Depression Scale (CES-D)
- **metacognition of allostatic control** (*M*): Sum of the subscales 3 (not worrying) and 8 (trusting) of the Multidimensional Assessment of Interoceptive Awareness (MAIA_3,8_).

One important caveat is that, to our knowledge, there does not yet exist a measure that was specifically developed for the construct of *M* (metacognition of allostatic control, i.e. the feeling of being in control over one’s own bodily states). In this study, as a proxy measure, we use the sum of the subscales 3 and 8 of the MAIA questionnaire. These subscales reflect an individual’s tendency not to experience distress in response to bodily inputs signalling dyshomeostasis and to perceive the body as a safe place, respectively. The sum of these subscales was used in a previous study testing predictions from ASE theory^33^ and may currently represent the best approximation to *M* that is easily applied in practice.

### SCM of the ASE theory

An SCM^5,25^ over variables **X** = [*X*_1_, …, *X*_*n*_] comprises a set of structural equations and distributions of the noise variables (a formal definition of an SCM is provided in Appendix A1). The structural equations together with the noise distributions induce the observational distribution P_**X**_ as simultaneous solution to the structural equations^43^. In addition to the observational distribution, an SCM induces interventional distributions. Each intervention denotes a scenario in which we fix a certain subset of the variables to a certain value, e.g. 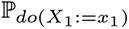.

Under assumptions of linearity and normality, the SCM of the ASE theory takes the following form:

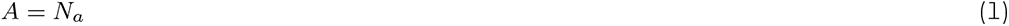

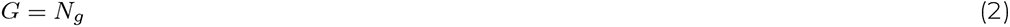

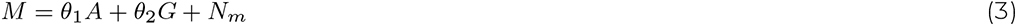

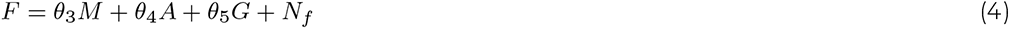

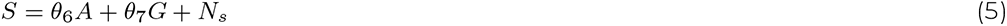

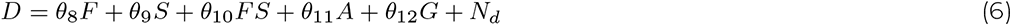

where *A* stands for age, *G* for gender, *M* for metacognition of allostatic control, *F* for fatigue, *S* general self-efficacy, *D* for depression, and *N*_*i*_ are jointly independent noise variables. ∀*i* ≠ *g, N*_*i*_ follows a normal distribution and *N*_*g*_ is a Bernoulli random variable.

Figure 1 displays a graphical summary of the causal structure implied by the ASE theory in the form of a directed acyclic graph (DAG) *J*_0_. The directed edge from metacognition of allostatic control (*M*) to fatigue (*F*) represents the prediction that fatigue arises as a consequence of a metacognitive diagnosis by the brain – i.e. the brain concludes that it has low control over its bodily states. When this low allostatic self-efficacy (for which fatigue is the accompanying feeling state) is combined with beliefs of lack of control in other domains than the body (low general self-efficacy), this is predicted to lead to the onset of depression. These effects are represented by the directed edges from fatigue (*F*) to depression (*D*) and from general self-efficacy (*S*) to depression (*D*). The variables age (*A*) and gender (*G*) are not explicitly part of the ASE theory, but are known to be associated with the central quantities of the theory. Hence, the DAG *J*_0_ in Figure 1 is representative for the induced observational distribution ℙ and the interventional distributions induced by interventions on metacognition of allostatic control (*M*; ℙ _*do*(*M* :=*m*)_), fatigue (*F*; ℙ _*do*(*F* :=*f*)_) or general self-efficacy (*S*; ℙ _*do*(*S*:=*s*)_).

**Figure 1:**
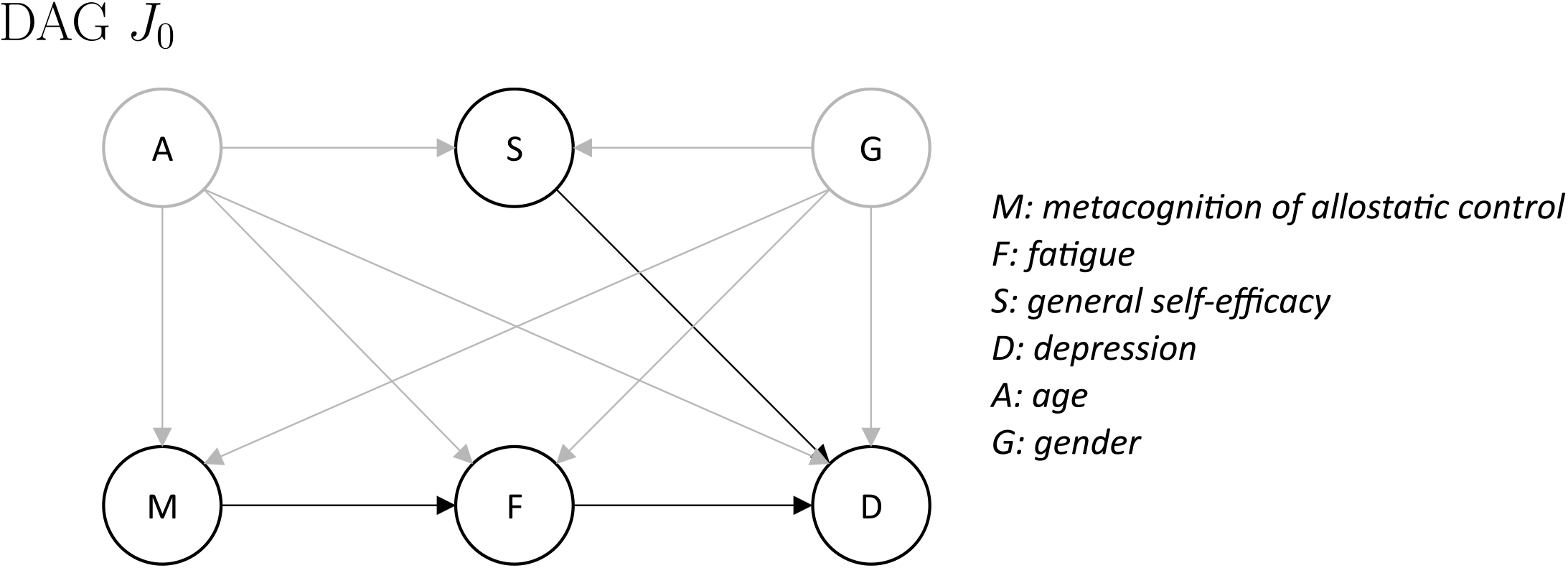
Directed acyclic graph (DAG) *J*_0_ summarizing the key proposal of the allostatic self-efficacy theory (ASE;^40^). The DAG *J*_0_ is representative for the induced observational distribution ℙ and the interventional distributions induced by interventions on metacognition of allostatic control (*M*; ℙ _*do*(*M*:=*m*)_), fatigue (*F*; ℙ _*do*(*F* :=*f*)_) or general self-efficacy (*S*; ℙ _*do*(*S*:=*s*)_). The other variables in the graph are depression (*D*), age (*A*) and gender (*G*). Black edges represent causal directions as proposed by the ASE theory, grey edges represent effects that are not explicitly part of the ASE theory but are likely to exist.

When taking a closer look at the causal graph in Figure 1, there are a number of points worth highlighting. (i) There is no direct link between metacognition of allostatic control (*M*) and general self-efficacy (*S*). (ii) There is no direct link from metacognition of allostatic control (*M*) to depression (*D*). All of its influence is mediated by fatigue (*F*). (iii) There is no direct link between fatigue (*F*) and general self-efficacy (*S*). While these three links are, in principle, plausible causal influences, they were not included in the original formulation of the ASE theory^40^. Whether these links should be included in a revision of the ASE theory can, in principle, be tested using methods of causal inference, given appropriate readouts of the involved quantities and relying on the assumption of the Markov condition.

### Statistical Analysis

Our hypotheses as well as the entire analysis were pre-registered in a time-stamped analysis plan that is publicly available on the Zenodo open data repository at https://doi.org/10.5281/zenodo.10559656. Below, we explicitly highlight any deviations from the pre-specified analysis plan. The analysis code is available at https://github.com/alexjhess/pbihb-ase-causality. The analysis pipeline underwent an internal code review by a researcher not involved in the initial data analysis to identify errors and ensure the reproducibility of our results.

### Causal structure of ASE theory in the PBIHB dataset

Learning causal structure from observational data is inherently difficult. One reason is the existence of models that are observationally but not interventionally equivalent^6,25,26,43^. This has several implications (e.g. see^43^), one of them being that without assumptions, it is impossible to learn causal structure from observational data.

In graphical models, the Markov condition (see e.g.^16^) is a formalisation of the following principle (sometimes referred to as Reichenbach’s common cause principle): If two random variables *X* and *Y* are dependent, then there must be some cause-effect structure that explains the observed dependence. That is, either *X* causes *Y*, or *Y* causes *X*, or another unobserved variable *H* causes both *X* and *Y*, or some combination of the aforementioned^29^. A formal definition of the Markov condition is presented in Appendix A2. The Markov condition establishes a connection from graphical separation properties (*d*-separation; see Appendix A3 for a formal definition) to conditional independencies in the distribution. Any distribution induced by an acyclic SCM satisfies the Markov condition with respect to the corresponding graph^17,25^. Hence, the Markov condition is typically considered to be a mild assumption.

Assuming that the observational distribution P induced by the SCM of the ASE theory (equations 1-6) is Markov with respect to the DAG *J*_0_, we tested whether we find any contradictions to the structure of the DAG *J*_0_ in the PBIHB dataset. More precisely, we examined the three predictions described in the last paragraph of and formalised as part of our pre-registered **Hypothesis 1:** Data from the PBIHB study satisfy the following conditional independence statements:

i. *M* ╨ *S* | *A*,*G*
ii. *M* ╨ *D* | *F*,*A*,*G* and *M* ╨ *D* | *F*,*A*,*G, S*
iii. *F* ╨ *S* | *A*,*G* and *F* ╨ *S* | *A*,*G*,*M*

As a statistical test for conditional independence, we used the asymptotic *χ*^2^ test on the mutual information for conditional Gaussians (MI_*cg*_) for mixed discrete and normal variables as implemented in the R package **bnlearn**^35^, using a significance level *α* = 0.01 (Bonferroni corrected).

Since conditional independence testing is a difficult statistical problem^38^, we validated our results using two alternative methods: a kernel conditional independence test (KCI;^45^) as implemented in the R package **CondIndTests**, and a test based on the generalised covariance measure (GCM;^38^) as implemented in the R package **GeneralisedCovarianceMeasure**. These additional tests of conditional independence were not part of our pre-specified analysis. We decided to conduct these additional tests to evaluate the robustness of our results across different methods of conditional independence testing (i.e. a sensitivity analysis). We used the same significance level *α* = 0.01 for the KCI as well as the GCM based tests to ensure compatibility with the pre-specified tests.

### Estimating the average causal effect from *M* to *F*

ASE theory predicts that fatigue is a feeling state that is triggered by a metacognitive diagnosis of loss of control over bodily states. We aimed to test this prediction as part of our **Hypothesis 2:** There is a negative average causal effect from metacognition of allostatic control (*M*) to fatigue (*F*)

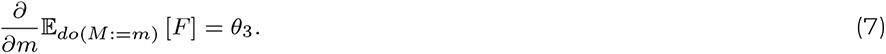

Adjusting for covariates is one of various methods for estimating causal effects from observational data. Suppose we are interested in finding the effect of *M* on *F* and assume the factors deemed relevant to the problem are structured as in Figure 1. In other words, we are interested in calculating the intervention distribution ℙ_*do*(*M* :=*m*)_(*f*). Given a valid adjustment set (VAS) **Z**, here e.g. **Z** = (*A, G*), the intervention distribution can be calculated (see^23,30,39^) as ℙ_*do*(*M* :=*m*)_(*f*) = ∑_*z*_ ℙ (*f* | *m*, **z**) ℙ (**z**), since

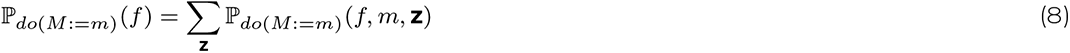

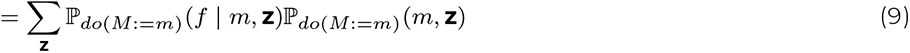

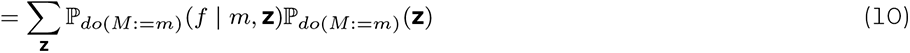

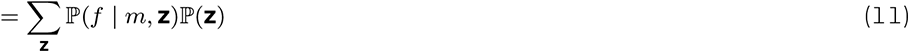

where in the last step one can use the fact that causal relationships are autonomous under interventions (this property is sometimes referred to as “autonomy”)^26^.

In linear Gaussian systems, a causal effect from *M* to *F* can be approximated by 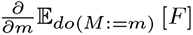 (see e.g.^26^). Assuming that **Z** is a VAS for *{M, F}* and *{M, F}*, **Z** follow a Gaussian distribution, then the conditional *F* | *M* = *m*, **Z** = **z** follows a Gaussian distribution as well. Hence, the mean of the distribution is given by

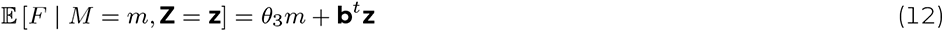

for some *θ*_3_ and **b**. It follows from equation 11 that

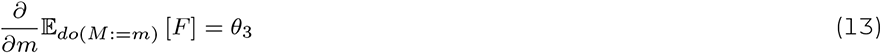

One can estimate the conditional mean (eq. 12) by regressing *F* on *M* and **Z** and subsequently reading off the regression coefficients for *M*. Alternatively, more sophisticated techniques for estimation of the average causal effect can be used, such as the propensity score method^32^ and double/debiased machine learning (DML;^8^). In Appendix B, the two methods are described in more detail.

As pre-specified in our analysis plan, we conducted linear regression in combination with a one-sided t-test on the regression coefficient of *M* to evaluate Hypothesis 2. We compared our estimate of the causal effect from *M* to *F* obtained via linear regression with the results obtained from using more sophisticated estimation techniques, i.e. the propensity score method^32^ and DML^8^, following our pre-registered analysis plan.

### Estimating the average causal effect from *F*\**S* on *D*

Another prediction of ASE theory is that fatigue, in combination with a generalisation of low self-efficacy beliefs beyond bodily control, induces depression. We formalised this prediction as part of our **Hypothesis 3:** There is a negative average causal effect of the interaction term between fatigue and general self-efficacy (*F*\**S*) on depression (*D*)

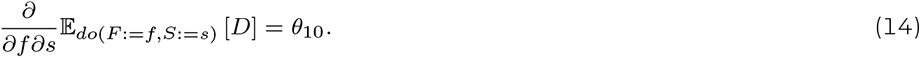

Evaluation of Hypothesis 3 followed the same line of reasoning as for Hypothesis 2. We used linear regression in combination with a one-sided t-test on the regression coefficient of *F*\**S*. Subsequently, we compared the resulting estimate to the results obtained using the propensity score method and DML.

## RESULTS

### Raw Data

Figure 2 shows a scatter plot matrix of the raw data. Displayed are the measures for all variables *A, G, M, F, S, D* used in the analysis.

**Figure 2:**
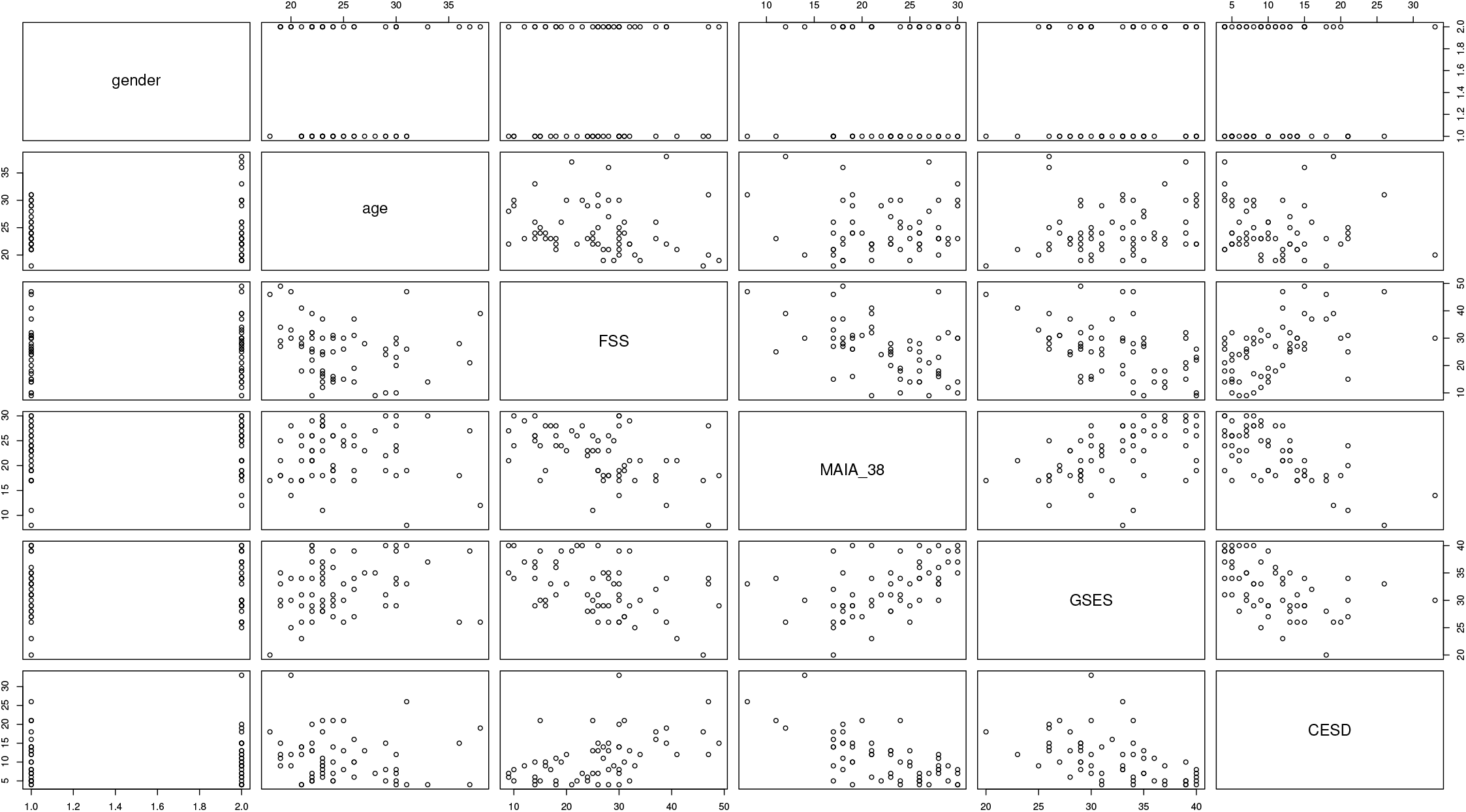
Scatter plot matrix of raw data used in the analysis. Displayed are all the pairwise scatter plots of the variables used for the analysis in a matrix format. For example, the scatter plot located on the intersection of row 3 and column 2 is a plot of variables age versus fatigue (as measured by the FSS). The variables displayed are gender, age, fatigue (assessed by the FSS), metacognition of allostatic control (assessed by the MAIA_3,8_), self-efficacy (assessed by the GSES) and depression (assessed by the CES-D).

### Results from the Statistical Analysis

#### Causal structure of ASE theory in the PBIHB dataset

Table 1 displays the results from conditional independence testing to evaluate the three predictions formulated as part of Hypothesis 1. The results can be summarised as follows:

**Table 1:**
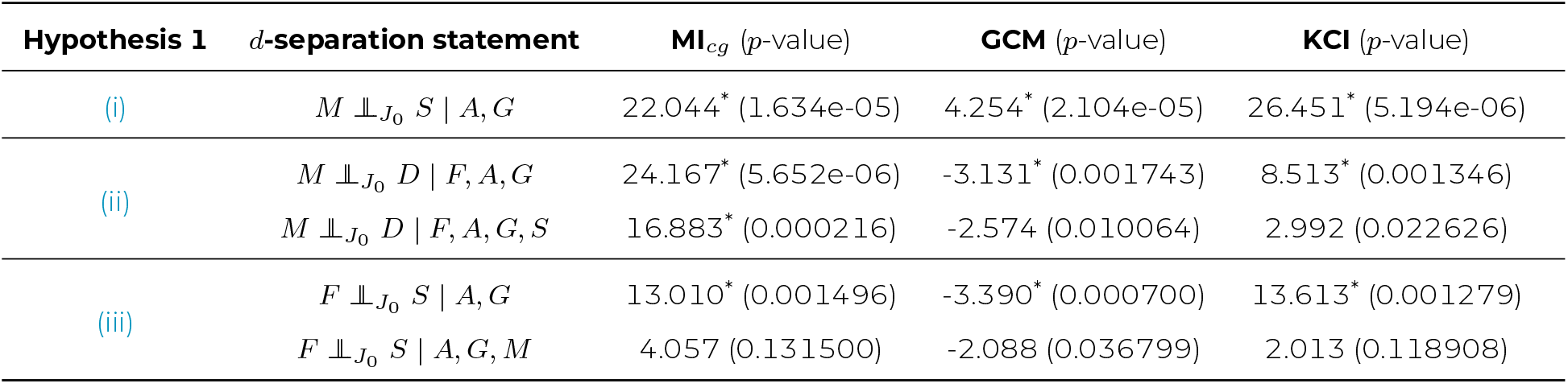
Results from different conditional independence test methods (MI_*cg*_, GCM, KCI) for the three predictions formulated as part of Hypothesis 1. Results are presented for three different test methods. An asterisk indicates statistically significant evidence against the null hypothesis (H_0_ : variables are conditionally independent) using the pre-specified level *α* = 0.01, which corresponds to a threshold of *p <* 0.05 Bonferroni corrected for the multiple comparisons of the five tests, *p*-values are shown in parentheses.

| Hypothesis 1 | $d$-separation statement | $MI_{cg}$ ($p$-value) | GCM ($p$-value) | KCI ($p$-value) |
| --- | --- | --- | --- | --- |
| (i) | $M \perp_{J_0} S \mid A, G$ | 22.044* (1.634e-05) | 4.254* (2.104e-05) | 26.451* (5.194e-06) |
| (ii) | $M \perp_{J_0} D \mid F, A, G$ | 24.167* (5.652e-06) | -3.131* (0.001743) | 8.513* (0.001346) |
| | $M \perp_{J_0} D \mid F, A, G, S$ | 16.883* (0.000216) | -2.574 (0.010064) | 2.992 (0.022626) |
| (iii) | $F \perp_{J_0} S \mid A, G$ | 13.010* (0.001496) | -3.390* (0.000700) | 13.613* (0.001279) |
| | $F \perp_{J_0} S \mid A, G, M$ | 4.057 (0.131500) | -2.088 (0.036799) | 2.013 (0.118908) |

i. 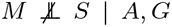. We find significant evidence that metacognition of allostatic control (*M*) and general self-efficacy (*S*) are not independent conditional on age (*A*) and gender (*G*) across all three different conditional independence test methods. In other words, we find a contradiction regarding the conditional independence of *M* and *S* given *A, G*, within the DAG *J*_0_.
ii. 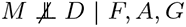 and *M* ╨ *D* | *F, A, G, S*. We find significant evidence that *M* and depression (*D*) are not independent conditional on fatigue (*F*), *A, G* across all three methods for conditional independence testing. This result is consistent with our findings for (i) in the sense that if we add a directed edge from *M* to *S* in the DAG *J*_0_ (Figure 1), the only set of variables that *d*-separates *M* and *D* is the set *F, A, G, S* (and not *F, A, G*). However, the results for conditional independence tests of *M* and *D* conditional on *F, A, G, S* are mixed with 2 out of 3 tests (GCM and KCI) not reaching the pre-specified significance level *α* = 0.01. Hence further evidence is needed to draw conclusions regarding the statement *M* ╨ *D* | *F, A, G, S*.
iii. 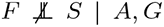 and *F* ╨ *S* | *A, G, M*. When looking at the conditional independence between *F* and *S*, the results depend on the set of variables that we condition on. We find significant evidence that *F* and *S* are not independent conditional on *A, G* across all three different test methods. However, we fail to reject the null hypothesis that *F* and *S* are independent conditional on the set *M, A, G* consistently across all three different test methods. This result is also in line with our findings for (i) in the sense that if we add a directed edge from *M* to *S* in the DAG *J*_0_ (Figure 1), the only set of variables that *d*-separates *F* and *S* is the set *M, A, G*.

#### Estimating the average causal effect from *M* to *F*

As predicted by the ASE theory, we find significant evidence for a negative average causal effect from metacognition of allostatic control (*M*) to fatigue 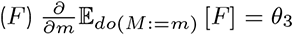 across all three different estimation methods. The resulting estimates 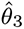 for the VAS **Z** = (*A, G*) are displayed in Table 2 alongside lower and upper bounds of a 95% confidence interval for 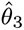, the corresponding value of the *t*-statistic as well as the *p*-value for the one-sided *t*-test.

**Table 2:** **Average causal effect from** *M* **to** *F* using **Z** = (*A, G*). Displayed are estimates of the average causal effect from *M* to 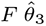 across three different methods to adjust for the covariates **Z** = (*A, G*). We report a point estimate 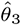, the lower and upper bounds of a 95% confidence interval for 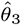, the value of the *t*-statistic as well as the *p*-value for the one-sided *t*-test. An asterisk indicates a statistical significance using the pre-specified level *α* = 0.017 (Bonferroni-corrected).

| estimation method | $\hat{\theta}_3$ | confidence interval | | $t$ value | $p$-value |
| --- | --- | --- | --- | --- | --- |
| linear regression | -0.4845* | -0.712 | -0.257 | -4.259 | 3.968e-05 |
| propensity score | -0.4816* | -0.717 | -0.246 | -4.092 | 6.689e-05 |
| DML | -0.3872* | -0.6481 | -0.1262 | -2.9082 | 0.0018 |

The results from our sensitivity analysis, i.e. estimating *θ*_3_ using a different VAS Z = (*A, G, S*), are listed in Table 3. They confirm the finding of a negative average causal effect from *M* to *F* when using Z = (*A, G*) as a VAS. The main difference between the results of the two analyses are that the second analysis using Z = (*A, G, S*) yields a slightly lower absolute value for 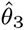 as well as a non-significant *p*-value using the DML method.

**Table 3:**
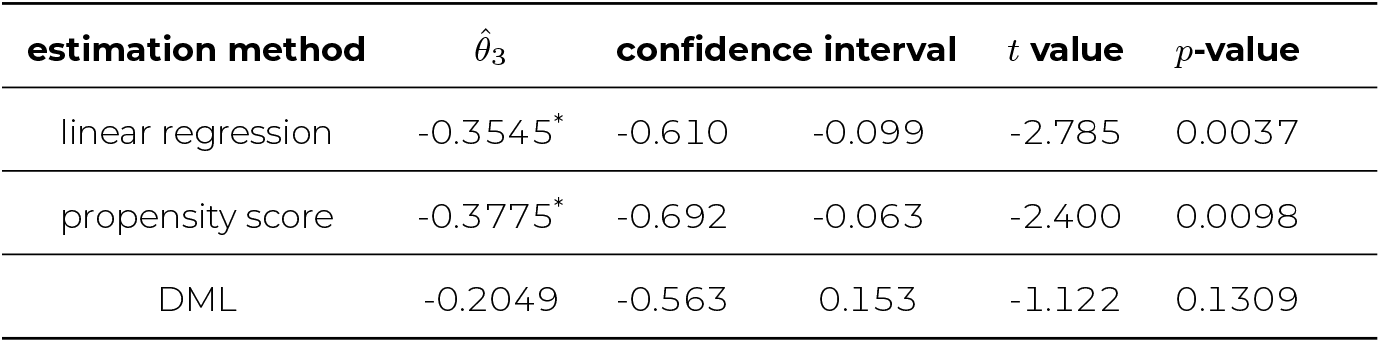
**Average causal effect from** *M* **to** *F* **using Z** = (*A, G, S*). Displayed are estimates of the average causal effect from *M* to 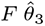 across three different methods to adjust for the covariates **Z** = (*A, G, S*). We report a point estimate of 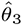, the lower and upper bounds of a 95% confidence interval for 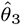, the value of the *t*-statistic as well as the *p*-value for the one-sided *t*-test. An asterisk indicates a statistical significance using the pre-specified level *α* = 0.017 (Bonferroni-corrected).

| estimation method | $\hat{\theta}_3$ | confidence interval | | $t$ value | $p$-value |
| --- | --- | --- | --- | --- | --- |
| linear regression | -0.3545* | -0.610 | -0.099 | -2.785 | 0.0037 |
| propensity score | -0.3775* | -0.692 | -0.063 | -2.400 | 0.0098 |
| DML | -0.2049 | -0.563 | 0.153 | -1.122 | 0.1309 |

**Table 4:** **Average causal effect of the interaction term** *F*\**S* **to** *D* **using Z** = (*A, G*). Displayed are estimates of the average causal effect of the interaction term *F*\**S* to *D* 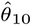 across three different methods to adjust for the covariates **Z** = (*A, G*). We report a point estimate of 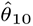, the lower and upper bounds of a 95% confidence interval for 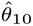, the value of the *t*-statistic as well as the *p*-value for the one-sided *t*-test. An asterisk indicates a statistical significance using the pre-specified level *α* = 0.017 (Bonferroni-corrected).

| estimation method | $\hat{\theta}_{10}$ | confidence interval | | $t$ value | $p$-value |
| --- | --- | --- | --- | --- | --- |
| linear regression | 0.0281 | -0.191 | 0.247 | 0.257 | 0.6010 |
| propensity score | 0.0142 | -0.151 | 0.180 | 0.172 | 0.5680 |
| DML | -0.2051 | -0.476 | 0.066 | -1.482 | 0.0691 |

**Table 5:** **Average causal effect of the interaction term** *F*\**S* **to** *D* **using Z** = (*A, G, S*). Displayed are estimates of the average causal effect of the interaction term *F*\**S* to 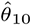 across three different methods to adjust for the covariates **Z** = (*A, G, M*). We report a point estimate of 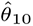, the lower and upper bounds of a 95% confidence interval for 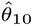, the value of the *t*-statistic as well as the *p*-value for the one-sided *t*-test. An asterisk indicates a statistical significance using the pre-specified level *α* = 0.017 (Bonferroni-corrected).

| estimation method | $\hat{\theta}_{10}$ | confidence interval | | $t$ value | $p$-value |
| --- | --- | --- | --- | --- | --- |
| linear regression | 0.0671 | -0.122 | 0.257 | 0.711 | 0.7599 |
| propensity score | 0.0337 | -0.159 | 0.227 | 0.350 | 0.6362 |
| DML | 0.0153 | -0.283 | 0.314 | 0.100 | 0.5399 |

#### Estimating the average causal effect from *F*\**S* to *D*

We do not find evidence for the predicted negative average causal effect of the interaction term between fatigue and general selfefficacy (*F*\**S*) on depression 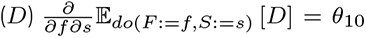 across all three different estimation methods for both VAS **Z** = (*A, G*) and **Z** = (*A, G, M*). Tables containing the resulting estimates for 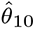 including a 95% confidence interval and the value of the *t*-statistic as well as the *p*-value for the one-sided *t*-test are listed in the Appendix C.

## DISCUSSION

In this paper, we proposed a formulation of the allostatic self-efficacy (ASE) theory of fatigue and depression in the language of causal inference. Specifically, we identified the variables of central interest to the ASE theory and formulated a structural causal model (SCM) under assumptions of linearity and normality. The SCM as well as the induced directed acyclic graph (DAG) describe the direction of causality among these variables. Using data of 60 healthy individuals from a previous study on interoception of breathing and its relation with several psychopathological constructs^14^, we tested the proposed causal model empirically. Relying on the assumption of the Markov condition, we used the dataset to search for contradictions to conditional independence statements (Hypothesis 1) that are implied by the graph structure (*d*-separation). In a second and third step, we estimated the value of two causal effects that are predicted by the ASE theory using methods of covariate adjustment, propensity scores and double/debiased machine learning. As predicted by the ASE theory, we found a statistically significant negative average causal effect from metacognition of allostatic control (*M*) to fatigue 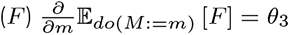 across all three methods of estimation. Our sensitivity analysis using a different valid adjustment set largely confirmed this finding with two out of three estimation methods yielding a significant result.

The assumption of the Markov condition establishes a connection from *d*-separation statements in a causal graph to conditional independence statements in the distribution. In the analysis of Hypothesis 1, we tested concrete predictions implied by the DAG *J*_0_ (Figure 1). (i) Using the the data from the PBIHB study, we were able to reject the null hypothesis of *M* ╨ *S* | *A, G* at the prespecified level *α* = 0.01. (ii) We found significant evidence against *M* ╨ *D* | *F, A, G* in the empirical data set. However, in line with the graph structure *J*_0_ implied by the ASE theory, we did not find clear evidence against *M* ╨ *D* | *F, A, G, S*. That is, only one out of three conditional independence tests rejected the null hypothesis of metacognition of allostatic control (*M*) being independent from depression (*D*) conditional on the set *F, A, G, S*. (iii) We also found significant evidence against *F* ╨ *S* | *A, G* in the empirical data. Yet, we did not find any evidence against (iii) *F* ╨ *S* | *A, G, M*. All three conditional independence test methods consistently failed to reject the null hypothesis of fatigue (*F*) and general self-efficacy (*S*) being independent given the set *A, G, D, M*. There are a number of potential explanations for the results related to Hypothesis 1. The most straightforward explanation is that the proposed causal model is incorrect. This can include the presence of additional edges between nodes as well as variables that were not considered acting as mediators or confounds or a combination of all of the aforementioned. For example, although the ASE theory does not make an explicit statement about a direct link between metacognition of allostatic control (*M*) and general self-efficacy (*S*), it is plausible to assume the existence of a directed edge from *M* (the feeling of control over bodily states) to *S* (an individuals general expectation of personal mastery and control^4^). The construct of *S* is closely related to concepts of metacognition (see e.g.^9^) and represents a “global” construct of self-beliefs about one’s capacity to achieve goals and overcome adversity; this can be understood as including more “local” domain-specific forms of self-efficacy, such as metacognition of allostatic control. From this view, the idea that metacognition of allostatic control (*M*) may contribute to (an thus influence) beliefs of general selfefficacy (*S*) is therefore not entirely unreasonable and would be a potential explanation for the results of (i) and (iii). More precisely, a directed edge from *M* to *S* would render *M* and *S d*-connected, since there would always exist a path between *M* and *S* that is not blocked by any set of variables. This cause-effect structure would explain the observed dependence between the two variables in the empirical data set according to Reichenbach’s common cause principle^29^. Another consequence of introducing and edge from *M* to *S* would be that the set of variables that *d*-separates *F* and *S* would consist of variables *A, G, M* and not *A, G* only, which corresponds to our findings for (iii). The same is true for the set of variables *d*-separating *M* and *D*, which would consist of variables *F, A, G, S* and not *F, A, G* in this case, potentially explaining our findings for (ii). However, since the evidence for (ii) *M* ╨ *D* | *F, A, G, S* was mixed, further research needs to bring clarity to the question of (conditional) independence of *M* and *D*.

The revised DAG *J*_1_ (Figure 3) provides a graphical summary of the above considerations regarding the results related to Hypothesis 1. From DAG *J*_0_ to *J*_1_, we added a directed edge from *M* to *S*. However, there are several other potential explanations for the observed results, so this example should by no means be taken as “the correct model”. If anything, this should be regarded as an updated hypothesis to be tested in future investigations.

**Figure 3:**
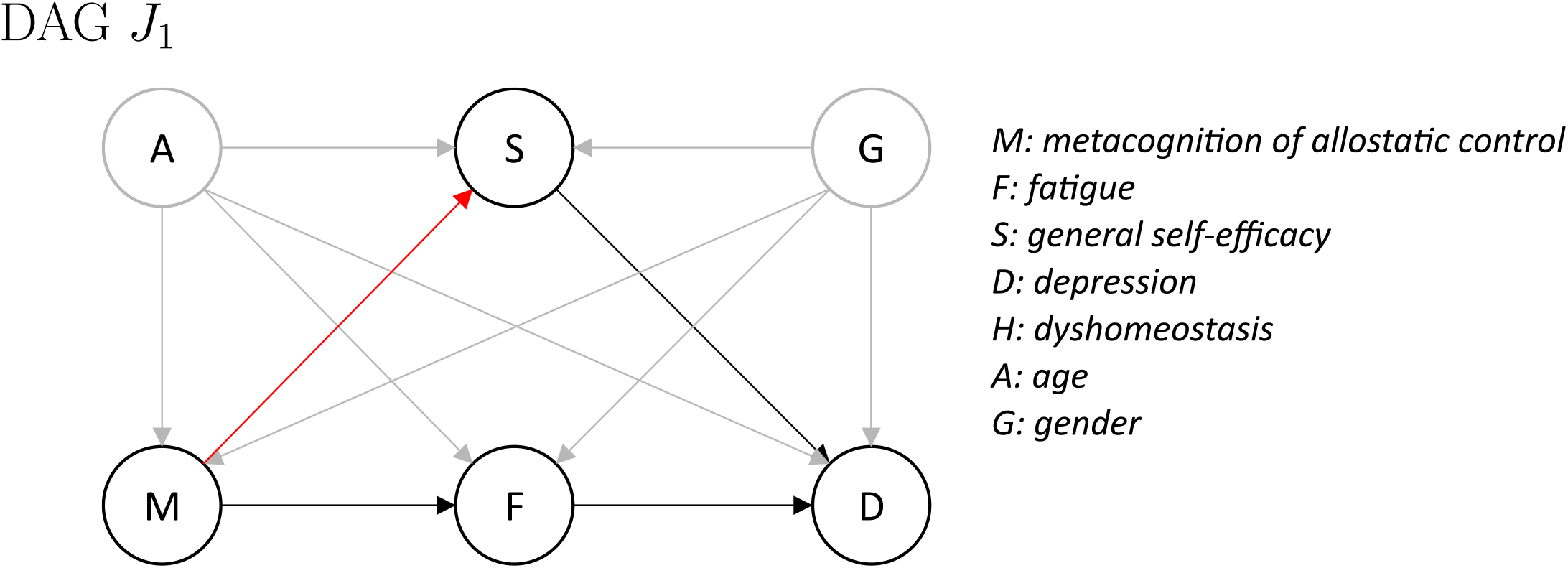
Updated directed acyclic graph (DAG) *J*_1_ of the allostatic self-efficacy theory (ASE;^40^) providing one potential explanation for the observed results from analysis of Hypothesis 1. Modifications from DAG *J*_0_ to *J*_1_ are shown in red.

Concerning Hypothesis 2, we found evidence for a negative average causal effect from metacognition of allostatic control (M) to fatigue 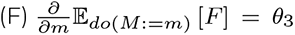 across all three estimation methods (covariate adjustment, propensity scores, DML) for two different VAS. This is in line with the prediction by the ASE theory that the subjective experience of fatigue arises as a consequence of a metacognitive diagnosis that the brain’s control over bodily states is failing (low allostatic control). This also confirms findings from previous research, which identified metacognition of allostatic control (*M*) (operationalised by the sum of the subscales 3 and 8 of the MAIA questionnaire) to be associated with fatigue (*F*) scores^33^. Our new results go beyond this previous finding, in the sense that the current study suggests a direction of the effect as opposed to purely associative statements. It is worth highlighting that the estimation of the causal effect from *M* to *F* would not be affected by the proposed additional link between *M* to *S* as suggested by the analysis results concerning Hypothesis 1 (i) (see Figure 3) since the set *A, G* would still be a valid adjustment set (VAS). With regard to Hypothesis 3, we did not find evidence for a negative average causal effect of the interaction term between fatigue and general self-efficacy (*F*\**S*) on depression 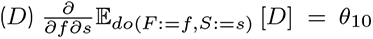. The present work is, to the best of our knowledge, the first attempt to investigate the predicted influence of the interaction between fatigue and general self-efficacy on depression. Across all three different estimation methods and using different VAS, we found, if anything, very small effects. However, one may rightfully question whether the sample in this study was adequate for testing Hypothesis 3, at least in the context of the ASE theory. This is because our participants were drawn from the general population and, not surprisingly, did not show pronounced levels of depression (compare Figure 2). By contrast, predictions of the ASE theory concerning depression assume a clinically relevant state of depression^40^. Therefore, the potential interaction effect *F*\**S* on *D* remains an open question that should be addressed in the future, using samples with clinically relevant levels of depression.

The present study has a number of limitations. First of all, we are limited by certain features of the dataset at hand. In addition to the low levels of depression discussed above, the sample size (N=60) is relatively small. Therefore, it will be crucial to see whether our findings can be reproduced in larger population samples. Moreover, the dataset is purely observational, meaning that there are no interventions on any of the variables of interest. This makes the problem of causal inference (even more) challenging. A logical aim for future studies would be to use variables like *M* as targets for cognitive interventions.

Additionally, our analysis relies on the assumption that we have access to valid measurements of the variables in our SCM. While we employed validated and widely used measures for fatigue, depression, and general self-efficacy, there does not yet exist a val-idated measurement tool that was specifically developed for the construct of metacognition of allostatic control (*M*). Here, as in previous research^33^, we used a plausible proxy measure, the sum of the MAIA subscales 3 and 8 (for a detailed motivation, please see the Methods section). An important goal for future research is the development and validation of easily applicable readouts for metacognition of allostatic control (*M*).

Beyond the limitations of the dataset, our proposed SCM of the ASE theory is arguably only a crude approximation to reality. The most obvious concern is the one of unobserved confounds, which we articulated in more detail in the discussion of the results from Hypothesis 1. More specifically, one important limitation of the present study is that our SCM does not include sleep. While sleep is not an explicit component of the ASE theory, previous work has repeatedly demonstrated the importance of sleep quality for fatigue (e.g.^19,33^). In the present study, we did not examine the potential influence of sleep since the available dataset did not include any measures of sleep quality.

A second limitation is that we adopt the common assumption that all effects are linear and that all of the random variables follow a normal distribution (except gender). These assumptions of linearity and normality should be kept in mind when interpreting our findings for Hypotheses 2 and 3. Another potential drawback of our SCM is that we did not explicitly consider the role of time. Most of the variables in our SCM are plausibly considered to be dynamic states, i.e. their values are likely to change over time. In this work, we used a dataset representing a snapshot in time and implicitly assumed that the causal effects take place instantaneously. However, it is plausible to assume that, for example, the effects of elevated fatigue levels do not immediately lead to elevated symptoms of depression, but that this effect evolves over timescales of weeks, months or even years.

Finally, there are numerous assumptions underlying our statistical tests. Conditional independence testing, which lies at the heart of causal discovery^39^, is one of its most challenging tasks^34^. For Hypothesis 1, we additionally rely on the assumption of the Markov condition. Without going into details for any of these assumptions, we highlight that the strongest of all the assumptions made throughout the entire analysis is the assumption of unconfoundedness. In other words, our results are based on the assumption that our proposed SCM contains all variables relevant for the phenomenon under consideration. However, it is likely that further variables exist that influence those in the proposed SCM (e.g. sleep, see above). The omission of these (partially unknown) variables may affect the results for all three hypotheses that we tested.

Despite the numerous limitations, this work also has several strengths worth highlighting. Foremost, we provided the first concrete formulation of the ASE theory in the language of causal inference. Our proposal of an SCM brings the content of a verbally formulated theory into the realm of concrete mathematical equations. Together with the induced DAG, they provide a formal basis for analysis and allowed us to identify a set of empirically testable hypotheses which may guide future research. Secondly, we used multiple independent methods for both conditional independence testing (Hypothesis 1) as well as the estimation of causal effects (Hypotheses 2 and 3). In this way, we are able to draw conclusions in that they do not depend on assumptions and properties of any single method. Last but not least, all of our hypotheses and statistical analysis procedures were pre-registered and specified in detail in an ex ante analysis plan (https://doi.org/10.5281/zenodo.10559656). Preregistration is an important and effective protection for the robustness of research, given the many degrees of freedom and the numerous cognitive biases that scientists may inadvertently be affected by^21^.

## CONCLUSIONS

In summary, our work provides a formal basis for testing predictions by the ASE theory of fatigue and depression in the context of causal inference. We evaluated central aspects of our proposed SCM using a publicly available dataset and provided an updated version of the SCM that accounts for our empirical findings. In addition, we were able to confirm previous findings regarding the association between metacognition of allostatic control (*M*) and fatigue (*F*). Our analysis enabled us to quantify the direction as well as the sign of the causal effect, i.e. we found a negative average causal effect from *M* to 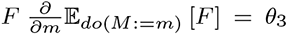, as predicted by the ASE theory. Finally, we identified a number of open questions that remain to be addressed in future research and that may help unravel the mechanisms behind fatigue and depression.

## Data Availability

https://doi.org/10.5281/zenodo.10992529

## ACKNOWLEDGMENTS

We wish to thank Jonas Peters for helpful discussions.

## AUTHOR CONTRIBUTIONS

Conceptualization, A.J.H., D.W., J.H. and K.E.S.; methodology, A.J.H., J.H. and K.E.S.; software, A.J.H. and D.W.; validation, D.W.; formal analysis, A.J.H.; investigation, A.J.H. and O.K.H.; resources, O.K.H. and K.E.S.; data curation, O.K.H.; writing—original draft preparation, A.J.H.; writing—review and editing, A.J.H., D.W., O.K.H., J.H. and K.E.S.; visualization, A.J.H.; supervision, K.E.S.; project administration, A.J.H.; funding acquisition, O.K.H. and K.E.S. All authors have read and agreed to the published version of the manuscript.

## FUNDING

This research was funded by the René and Susanne Braginsky Foundation, the ETH Foundation and the University of Zurich. O.K.H. (née Faull) was supported by a Marie Sklodowska-Curie Postdoctoral Fellowship from the European Unions Horizon 2020 research and innovation program under the grant agreement 793580, and a Rutherford Discovery Fellowship from the Royal Society Te Aprangi.

## AUTHOR COMPETING INTERESTS

The authors declare no conflicts of interest.

## AI ASSISTED TECHNOLOGIES

We did not use any AI assisted technologies, neither for data analysis nor during writing of the manuscript.

## APPENDIX A. DEFINITIONS

### A1. Structural Causal Model

We adopt the definition of SCMs according to^43^:

#### Definition 1.

*An SCM over variables* ***X*** = [*X*_1_, …, *X*_*n*_] *comprises*

i. *structural equations which relate each variable X*_*k*_ *to its parents* ***PA***(*X*_*k*_) ⊆ *{X*_1_, …, *X*_*n*_*} and a noise variable N*_*k*_ *via a function f*_*k*_ *such that X*_*k*_ := *f*_*k*_ (***PA*** (*X*_*k*_), *N*_*k*_), *as well as a*
ii. *noise distribution* P_*N*_ *of the noise variables* ***N*** = [*N*_1_, …, *N*_*n*_]^*T*^.

*In a directed causal graph associated with an SCM, the nodes correspond to the variables X*_1_, …, *X*_*n*_ *and there is an edge from X*_*i*_ *to X*_*j*_ *whenever X*_*i*_ *appears on the right hand side of the equation X*_*j*_ := *f*_*j*_ (***PA*** (*X*_*j*_), *N*_*j*_). *In other words, if X*_*i*_ ∈ ***PA***(*X*_*j*_) *the graph contains the edge X*_*i*_ → *X*_*j*_. *For this work, we assume that the graph does not contain any cycles. The structural equations together with the noise distributions induce the observational distribution* P_*X*_ *of X*_1_, …, *X*_*n*_ *as simultaneous solution to the equations*.

### A2. Markov condition

#### Definition 2.

*Given a DAG G over nodes* ***X***, *we say that the distribution* P_***X***_ *satisfies*

i. *the* ***global Markov property*** *(MP) with respect to G if* ∀ *disjoint A, B, C* ⊆ ***X*** *A d-sep B* | *C* =⇒ *A* ╨ *B* | *C*
ii. *the* ***local Markov property*** *(MP) if* ∀*j X*_*j*_ ╨ *ND*_*j*_ |*PA*_*j*_
iii. *the* ***factorisation property*** *if* P_***X***_ *is absolutely constant with respect to a product measure and* 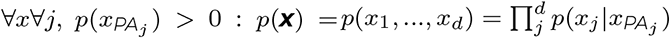

In the above definition, we used the following notation: ND_*j*_ represent the non-descendants of node *X*_*j*_ and PA_*j*_ denotes all nodes that have a directed edge to node *X*_*j*_.

### A3. *d*-separation

#### Definition 3.

*d-separation is a graphical criterion whether two nodes are connected or not. Let* ***X, Y, Z*** *disjoint*.

i. *A path X* = *i*_1_, …, *i*_*m*_ = *Y is blocked by* ***Z*** *⟺* ∃ *node i*_*k*_ *with i*_*k*−1_ → *i*_*k*_ → *i*_*k*+1_ *and i*_*k*_ ∈ ***Z*** *OR* ∃ *node i*_*k*_ *with i*_*k*−1_ ← *i*_*k*_ ← *i*_*k*+1_ *and i*_*k*_ ∈ ***Z*** *OR* ∃ *node i*_*k*_ *with i*_*k*−1_ ← *i*_*k*_ → *i*_*k*+1_ *and i*_*k*_ ∈ ***Z*** *OR* ∃ *node i*_*k*_ *with i*_*k*−1_ → *i*_*k*_ ← *i*_*k*+1_ *and i*_*k*_ ∈*/* ***Z*** *and DE*(*i*_*k*_) ∩ ***Z*** = ∅
ii. ***X, Y*** *are d-connected given* ***Z*** *⟺* ∃*X* ∈ ***X***, *Y* ∈ ***Y*** *s*.*t*. ∃ *path between X and Y that is not blocked*
iii. *if* ***X, Y*** *are not d-connected, then they are d-separated. We sometimes write* ***X*** *d-sep* ***Y*** | ***Z*** *or* ***X*** ╨_*G*_ ***Y*** | ***Z***

## APPENDIX B. ESTIMATING CAUSAL EFFECTS USING COVARIATE ADJUSTMENT

### B1. The “propensity score” method

In a point treatment situation one can adjust for a set of confounders **Z** = (*A, G*) when estimating the effect of exposure *M* by weighting observations *i* by the inverse probability weights

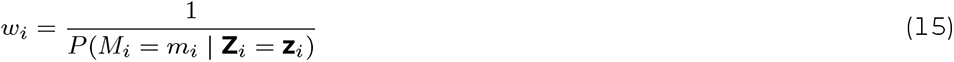

To increase statistical efficiency, one can use stabilised weights, e.g.

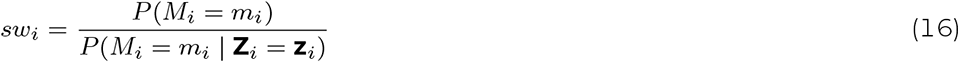

When dealing with a continuous exposure variable *M*, one can use stabilised weights

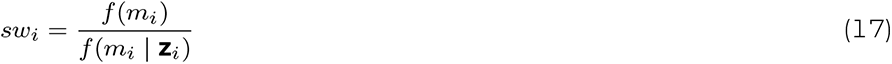

where *f* (*m*_*i*_) is the marginal density function of *M*, evaluated at the observed value in unit *i, m*_*i*_, and *f* (*m*_*i*_ | **z**_*i*_) conditional density function of *M* given **Z**, evaluated at the observed values in unit *i, {m*_*i*_, **z**_*i*_*}*^42^. Weighting observations *i* by *sw*_*i*_, one can fit a causal model, for instance a marginal structural model (MSM)

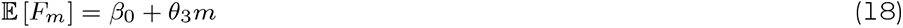

with continuous outcome fatigue *F*. The response variable *F*_*m*_ is the potential outcome that could have been observed in a unit under study, when that unit would have received a specific treatment level *m*^31^. The expectation E [*F*_*m*_] is the mean outcome, when all units under study would have received a specific treatment level *m*. Parameter *θ*_3_ then quantifies the causal effect of *M* on *F*^42^.

### B2. Double/Debiased Machine Learning

DML removes the impact of regularisation bias and overfitting on estimation of the parameter of interest *θ*_3_ by using Neyman-orthogonal moments and cross-fitting^8^. One application of DML is in the context of a partial linear regression model,

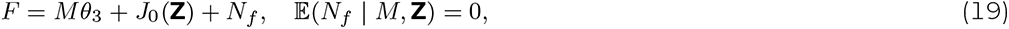

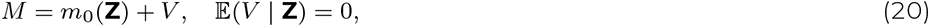

with fatigue *F*, metacognition of allostatic control *M*, a VAS Z = (*A, G*) consisting of confounding covariates and stochastic error terms *N*_*f*_ and *V*. The confounding covariates Z affect *M* and *F* via the functions *m*_0_ and *J*_0_, respectively. DML can be used to estimate *θ*_3_, i.e. the main regression coefficient that we would like to infer, which can be interpreted as the average causal effect from M to F^3^.

## APPENDIX C. RESULTS FROM ESTIMATING THE AVERAGE CAUSAL EFFECT FROM *F*\**S* TO *D*

## Notes

### Competing Interest Statement

The authors have declared no competing interest.

### Clinical Protocols

https://doi.org/10.5281/zenodo.10559656

https://github.com/alexjhess/pbihb-ase-causality

### Funding Statement

This research was funded by the Rene and Susanne Braginsky Foundation, the ETH Foundation and the University of Zurich. O.K.H. (nee Faull) was supported by a Marie Sklodowska-Curie Postdoctoral Fellowship from the European Union's Horizon 2020 research and innovation program under the grant agreement 793580, and a Rutherford Discovery Fellowship from the Royal Society Te Apārangi.

### Author Declarations

https://doi.org/10.5281/zenodo.10992529

